# Efficacy of Linear Regression Modelling of SARS-CoV-2 cases based on local wastewater surveillance

**DOI:** 10.1101/2022.10.20.22281303

**Authors:** Martin Lehmann, Michael Geissler, Waldemar Hahn, Richard Gebler, Björn Helm, Roger Dumke, Alexander Dalpke, Markus Wolfien

**Author notes:** Corresponding Author: Martin Lehmann. Contributed equally (Martin Lehmann, Michael Geissler).

## Abstract

In the ongoing SARS-CoV-2 pandemic, there is a need for new strategies for surveillance and identification of arising infection waves. Reported cases of new infections based on individual testing are soon deemed inaccurate due to ever changing regulations and limited testing capacity. Wastewater based epidemiology is one promising solution that can be broadly applied with low efforts in comparison to current large-scale testing of individuals. Here, we are combining local wastewater data from the city of Dresden (Germany) along with reported cases and vaccination data from a central database (Robert-Koch-Institute) with virus variant information to investigate the correlation of virus concentrations in the wastewater and reported SARS-CoV-2 cases. In particular, we compared Linear Regression and Machine Learning (ML) models, which are both revealing an existing correlation of virus particles in wastewater and reported cases. Our findings demonstrate that the different virus variants of concern (Alpha, Delta, BA.1, and BA.2) contribute differently over time and parameters vary between variants, as well. By comparing the Linear Regression and ML-based models, we observed that ML can achieve a good fit for training data, but Linear Regression is a more robust tool, especially for new virus variants. We hereby conclude that deriving the rate of new infections from local wastewater by applying Linear Regression may be a robust approximation of tracing the state of the pandemic for practitioners and policy makers alike.

## 1 Introduction

Recently, more contagious but less severe variants of the severe acute respiratory syndrome coronavirus 2 (SARS-CoV-2) occurred and encountered increasing vaccination rates. At the same time community test resources, as well as testing rigour, are slowly diminishing. Hence, fewer people may be tested, which poses the challenge of underestimating the community transmissions by magnitudes. This affects both political and clinical pandemic management, community trust, and general risk awareness. Since the beginning of the current pandemic, researchers have been studying the efficacy of methods to detect the presence of the novel SARS-CoV-2 virus in municipal wastewater treatment plants (WWTP). Wastewater-based epidemiology (WBE) shall be used as an early warning system since it shows changes in community transmission levels days before affected persons even recognise their infection and get themselves tested, if done at all (Olesen et al., 2021). In addition, long-term monitoring can be achieved at a low cost and low impact compared to individual testing approaches (Daughton, 2020; Han et al., 2022).

The current gold standard for the detection of the SARS-CoV-2 pathogen is the quantitative reverse transcriptase polymerase chain reaction (RT-qPCR) to target different genomes by means of specific primers. Advantages of RT-qPCR include high sensitivity and specificity, as well as wide adoption and availability. Disadvantages are mainly due to relatively high costs compared to other individual clinical testing methods (e.g., rapid antigen tests) and a long analysis time (Pérez-López & Mir, 2021). However, for wastewater surveillance, these disadvantages are negligible due to the low sampling frequency (once a day) and high pool-testing ratio (one test required for hundreds of thousands of people). RT-qPCR allows for additional quantification of the collected SARS-CoV-2 material (Nagura-Ikeda et al., 2020). All existing variants of concern (VOC) of this virus (e.g., Alpha, Delta) can be detected by means of RT-qPCR tests with a high sensitivity, except for one specific S-gene target failure. Therefore, the recommendation is to test for a minimum of two different genome targets (Ferré et al., 2022), which was also done in this study. Furthermore, since virus generation times (time of a complete infection cycle) and presence of distinct symptoms vary between variants and there remains a time lag upon official case reporting, it is expected that time lags between wastewater and reported cases may be different between virus variants as well (Abbott et al., 2022; Hart et al., 2022; Olesen et al., 2021). Following advances in sample treatment and analysis methods, test sensitivity appears high for these methods, so that SARS-CoV-2 residues can be detected in wastewater with a lower threshold of 13/100k people (Manuel et al., 2021).

However, interpretation and quantification of results of wastewater investigations is difficult in relation to officially reported community transmission levels since there are many differing influence factors on both approaches. The European Union (EU) is recommending the use of WBE even beyond the current pandemic for all its member states (Sinkevicius, 2021) and the World Health Organization (WHO) recognises its advantages alike (World Health Organization, 2020). Nevertheless, without a clear interpretation guideline for practitioners and policy makers, WBE alone might be prevented from delivering on its full potential. Hence, we are proposing a means of linking the results with the well-known and established cases of new infections towards specific virus variants.

This work shall investigate how effectively local wastewater data can be used to estimate community transmission levels (reported by the German authority “Robert Koch Institut” [<u>RKI</u>]) based on data from a medium-sized German city, namely Dresden (∼500,000 inhabitants, area of 330 km²), in the time from 15^th^ March 2021 to 30^th^ April 2022. Our study includes a comparison between Linear Regression and Deep Learning (DL) models with Long Short-Term Memory (LSTM) units, to account for an interdependency and precise estimation between locally reported SARS-CoV-2 incidence and virus level in the wastewater.

## 2 Methods

### 2.1 Dataset

For this analysis, SARS-CoV-2 concentrations in wastewater, reported case numbers, the virus variant distribution, vaccination status, as well as precipitation were combined into a daily time-series dataset. Cases are reported in numbers of infected people or in incidence rates (cases per 100,000 people). The latter allows a better comparison of regions with very different population densities. All of the above-mentioned data was sampled from five different data sources as the following:

i. The wastewater sampling, measurement and analysis were conducted and results were provided by the TU Dresden wastewater and virology research groups (M. Geissler, R. Dumke, A. Dalpke). The data comprises of daily virus concentration measurements from wastewater samples (discharge proportional 24h composite samples, collected after the grit chamber and before primary clarification) for the Dresden area (WWTP Dresden-Kaditz). In the present study, the raw wastewater samples were analysed by means of polyethylene glycol precipitation (PEG), RNA extraction, PCR inhibitor removal, and final RT-qPCR using a commercial kit for amplification of E and S gene of SARS-CoV-2 as published earlier (Dumke et al., 2021). Here, concentrations of SARS-CoV-2 in wastewater are referred to as wastewater data.
ii. The reported cases were provided by the RKI on their Github-page (https://github.com/robert-koch-institut/SARS-CoV-2_Infektionen_in_Deutschland). It contains data for each day and age group with an update of infection rates (also looking back in time in case of late reports or error corrections). After aggregation, one row remains for each day based on the most recent data. There were two different date columns available: (a) when the result was reported to the authorities, (b) when the patient reported symptom onset (which is usually a few days earlier; if no date was reported by the patient, it matches (a)). As reference for later analysis, the symptom onset date is used (“Refdate”), instead of the reporting date. Doing this will align the data better with the SARS-CoV-2 concentration in wastewater.
iii. RKI data for virus variant sequencing (https://github.com/robert-koch-institut/SARS-CoV-2-Sequenzdaten_aus_Deutschland) contains a table of analysed virus samples with attributes of date, found lineage, and sampling reason. According to their provided legend, sampling reasons “X” (unknown) and “N” (None) were excluded.
iv. Vaccination data were also provided by RKI on their Github page (https://github.com/robert-koch-institut/COVID-19-Impfungen_in_Deutschland). Data is grouped by day and age group. It contains the number of vaccinated people in each group and how many shots they have received so far. From this, time-series were built for both the relative population with full vaccination status and booster status.
v. Precipitation data for Dresden was downloaded from Meteostat (https://meteostat.net/de/place/de/dresden?s=D1051&t=2021-03-15/2022-04-30) and contains accumulated samples for each day from 15^th^ March 2021 to 30^th^ April 2022.

### 2.2 Data preprocessing

The RKI data was filtered for the locality “SK Dresden” and daily reported cases were accumulated as moving sum for n previous days (being 7, 14, and 28) respectively for each sample at time point *t*. Accumulation for seven days was selected in accordance with the standard RKI reporting scheme, 14 days as it is handled often internationally, and 28 days was used as accumulation period in some previous studies (Galani et al., 2022; Medema et al., 2020; Zhu et al., 2022). Cases are represented as incidence rates, i.e. the case numbers were divided by the population number. Additionally, accumulates were normalised to seven days (equation (1)). This makes the results more comparable (same value range) and improves later interpretation and application of the model.

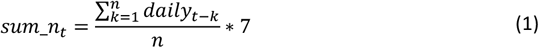

The wastewater data was also accumulated for 7, 14, and 28 days using equation (1) and will be analysed up until 30^th^ April 2022, allowing for another three weeks (as per 22^nd^ May 2022) of delayed case reporting (which is then attributed to an earlier date). All data was merged and gaps were linearly interpolated, as well as negative values excluded. In addition, for the linear model, all days with more than 5mm precipitation were excluded according to Rubio et al. (2021), then interpolated before aggregating additional columns.

The final data table summarises each variable together with their source and reason of inclusion after preprocessing (Table 1). A plot of the resulting time series is given in Figure 5.

**Table 1.** Summary of variables in the final dataset after preprocessing with data source and reason for inclusion. Data source numbering according to dataset section -wastewater.

| Variable name | Data Source | Reason for inclusion | Comment |
| --- | --- | --- | --- |
| WW daily | i (UKDD) | correlation parameter/<br>Machine Learning input | raw data |
| WW 7 days | i (UKDD) | correlation parameter/<br>Machine Learning input | aggregated 7 days |
| WW 14 days | i (UKDD) | correlation parameter/<br>Machine Learning input | aggregated 14 days |
| WW 28 days | i (UKDD) | correlation parameter/<br>Machine Learning input | aggregated 28 days |
| Cases daily | ii (RKI) | correlation parameter/<br>estimation target | raw data |
| Cases 7 days | ii (RKI) | correlation parameter/<br>estimation target | aggregated 7 days |
| Cases 14 days | ii (RKI) | correlation parameter/<br>estimation target | aggregated 14 days |
| Cases 28 days | ii (RKI) | correlation parameter/<br>estimation target | aggregated 28 days |
| Variant contribution: Alpha | iii (RKI) | linear model separation/<br>Machine Learning input | potential influence on case numbers |
| Variant contribution: Delta | iii (RKI) | linear model separation/<br>Machine Learning input | potential influence on case numbers |
| Variant contribution: BA.1 | iii (RKI) | linear model separation/<br>Machine Learning input | potential influence on case numbers |
| Variant contribution: BA.2 | iii (RKI) | linear model separation/<br>Machine Learning input | potential influence on case numbers |
| # People 2nd vaccination shot | iv (RKI) | Machine Learning input | potential influence on case numbers |
| # People 3rd vaccination shot | iv (RKI) | Machine Learning input | potential influence on case numbers |
| Precipitation daily | v (Meteostat) | preprocessing input/<br>Machine Learning input | potential dilution of wastewater |

### 2.3 Modelling of SARS-CoV-2 cases via Linear Regression

The simplest possible dependency between wastewater data and new infection cases is linear. Many studies found that correlation of confirmed cases and concentration of SARS-CoV-2 in wastewater can be very strong and models are effective (Ai et al., 2021; Medema et al., 2020). Under the assumption of linear correlation and because of the independence of scale of its results, we used Pearson correlation. In particular, the most common approach for time-series is to employ cross-correlation (Bracewell, 1965) and calculate a “lead time” or “lag” by which both wastewater and reported cases time series would need to be shifted against each other to yield a maximum correlation coefficient. Medema et al. (2020) suggest that community prevalence should be accumulated for 28 days (following indications that virus shedding continues for that long) and correlated against daily wastewater data. However, their data was from early 2020, only targeting the “wildtype” (unmutated/first occurring) virus variant and (comparably) low community transmission levels (Incidence < 100). Newer studies employ varying periods between 3 and 28 days of averaging depending on the target application. Interestingly, longer periods usually yield better correlation coefficients and can therefore be used when a single conversion factor is required (Galani et al., 2022; Weidhaas et al., 2021; Zhu et al., 2022).

A Linear Regression is fitted (using the Python library *”scikit-learn”*), which approximately represents the actual correlation between both series for each accumulation period, while accounting for the relative proportion of the respective variant at each point in time by weighting the sample contribution by equations (2) (sample weighting), (3) (weighted covariance), and (4) (weighted correlation coefficient). *i* is the sample index, *w*_*i*_ the variant specific contribution factor, *x*_*i*_ the sample value of the first time series and *y*_*i*_ the sample value of the other time series. Comparing actual and modelled cases, the (weighted) correlation coefficient (*r* ∈ [0; 1]) will be used as a performance metric for each partition and accumulation period, as well as for the entire data set.

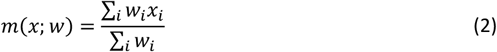

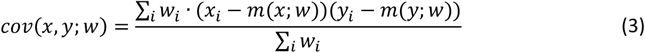

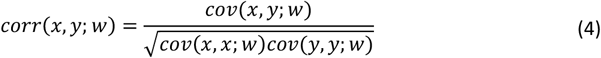

The virus variant data is filtered for variants of concern (VOC), rolling averaged with a window of ten days, to average out fluctuations in the sampling process (e.g. non-representative sampling, date not aligned with symptom onset), and the relative contribution to all samples is calculated.

### 2.4 Modelling of SARS-CoV-2 cases via Machine Learning

More complex interdependences of multiple variables could be described with non-linear models, such as neural networks. Machine Learning models have been trained to show an alternative route towards the estimation of actual infections. Sequential models with LSTM units have been successfully used in similar real-world applications (Ahmed et al., 2022; Nikparvar et al., 2021; Rashed et al., 2022).

The input data for this model comprises the wastewater data (accumulated for 14 days only, this seems a good compromise between smoothness and timeliness of the results), virus variant distribution, community vaccination status, and finally, the reported cases (14 days accumulated) as a target vector. All input data has been scaled to the target range of 0-1 and two different sequential models have been trained (Table 1), using Google’s python library “Tensorflow” (Abadi et al., 2016). Training data has been split into windows of constant length (60 samples each) and two sets of windows for training (75%) and validation (25%) of the model. A variable portion at the end of the dataset is excluded for final evaluation of the model’s performance (e.g. the BA.2 wave, see Figure 4). Therefore, the models are trained to predict each day’s actual reported cases based on the history of the input data summarised. The training was conducted on a free instance of Google’s Colaboratory platform using default GPU resources.

The properties of the layers of the models are outlined in Table 4 The first “simple” model has only 1 LSTM layer and ∼5,000 neurons, and serves as a simple baseline; the “deep” model comprises 4 LSTM layers and over 300,000 neurons to cover more complexity. LSTM units (“Long short-term memory”), introduced by Hochreiter & Schmidhuber (1997), are used to model recurrent patterns in time series. The main aspect is that non-linear correlations might also have a time dependent element of variable length (Staudemeyer & Morris, 2019). The size of the networks are based on literature findings and experimentation with the data. More details can be found in Table 4. Using variable-length windows, (covering the entire training data from the beginning) instead of fixed-length windows, has been considered for this work but did not yield better overall performance. The model is expected to learn the influence of wastewater virus load on the reported case numbers, which is a pattern that seems to repeat itself throughout the dataset with slightly different (and also time-dependent) parameters, such as vaccination status and currently dominant virus variant.

**Table 2.** Errors (RMSE) for all models, training and testing sections. Lowest errors for each column are in bold font.

|  | Linear Model |  |  |  |  |  |  | Simple LSTM Model |  |  |  |  |  |  | Deep LSTM Model |  |  |  |  |  |  |
| --- | --- | --- | --- | --- | --- | --- | --- | --- | --- | --- | --- | --- | --- | --- | --- | --- | --- | --- | --- | --- | --- |
| Train on→ | Alpha | Delta | BA.1 | BA.2 | Alpha<br>-><br>Delta | Alpha<br>-><br>BA.1 | Alpha<br>-><br>BA.2 | Alpha | Delta | BA.1 | BA.2 | Alpha<br>-><br>Delta | Alpha<br>-><br>BA.1 | Alpha<br>-><br>BA.2 | Alpha | Delta | BA.1 | BA.2 | Alpha<br>-><br>Delta | Alpha<br>-><br>BA.1 | Alpha<br>-><br>BA.2 |
| Test on ↓ |  |  |  |  |  |  |  |  |  |  |  |  |  |  |  |  |  |  |  |  |  |
| Alpha | **0.005** | 0.074 | 0.123 | **0.061** | 0.071 | 0.150 | 0.142 | **0.003** | 0.136 | 0.222 | **0.150** | **0.004** | **0.005** | **0.006** | **0.001** | 0.461 | 0.190 | 0.144 | **0.002** | **0.005** | **0.006** |
| Delta | 0.148 | **0.029** | 0.161 | 0.071 | **0.029** | 0.168 | **0.082** | 0.189 | **0.042** | 0.145 | 0.193 | 0.014 | 0.017 | 0.018 | 0.196 | 0.161 | 0.145 | **0.128** | 0.007 | 0.009 | 0.010 |
| BA.1 | 0.357 | 0.182 | **0.061** | 0.195 | 0.184 | **0.070** | 0.120 | 0.499 | 0.147 | **0.082** | 0.297 | 0.245 | 0.070 | 0.061 | 0.539 | **0.112** | **0.091** | 0.331 | 0.136 | 0.014 | 0.015 |
| BA.2 | 0.501 | 0.107 | 0.421 | 0.069 | 0.108 | 0.414 | 0.083 | 0.557 | 0.412 | 0.233 | 0.332 | 0.547 | 0.203 | 0.063 | 0.598 | 0.224 | 0.192 | 0.370 | 0.211 | 0.228 | 0.015 |
| Avg. | 0.253 | 0.098 | 0.191 | 0.099 | 0.098 | 0.201 | 0.107 | 0.312 | 0.184 | 0.171 | 0.243 | 0.203 | 0.074 | 0.037 | 0.334 | 0.239 | 0.155 | 0.243 | 0.089 | 0.064 | 0.012 |

**Table 3.** Pearson Correlation Coefficients for all models, training and testing sections. Lowest errors for each column are in bold font.

|  | Linear Model |  |  |  |  |  |  | Simple LSTM Model |  |  |  |  |  |  | Deep LSTM Model |  |  |  |  |  |  |
| --- | --- | --- | --- | --- | --- | --- | --- | --- | --- | --- | --- | --- | --- | --- | --- | --- | --- | --- | --- | --- | --- |
| Train on→ | Alpha | Delta | BA.1 | BA.2 | Alpha<br>-><br>Delta | Alpha<br>-><br>BA.1 | Alpha<br>-><br>BA.2 | Alpha | Delta | BA.1 | BA.2 | Alpha<br>-><br>Delta | Alpha<br>-><br>BA.1 | Alpha<br>-><br>BA.2 | Alpha | Delta | BA.1 | BA.2 | Alpha<br>-><br>Delta | Alpha<br>-><br>BA.1 | Alpha<br>-><br>BA.2 |
| Test on ↓ |  |  |  |  |  |  |  |  |  |  |  |  |  |  |  |  |  |  |  |  |  |
| Alpha | 0.975 | 0.975 | 0.975 | 0.975 | 0.975 | 0.975 | 0.975 | **0.988** | 0.830 | 0.772 | -0.553 | 0.985 | 0.983 | 0.982 | **0.999** | 0.737 | 0.681 | 0.915 | 0.995 | 0.997 | 0.996 |
| Delta | **0.985** | **0.985** | **0.985** | **0.985** | **0.985** | **0.985** | **0.985** | 0.929 | **0.977** | 0.598 | 0.630 | **0.997** | **0.995** | **0.994** | 0.800 | 0.614 | 0.868 | **0.960** | **0.999** | **0.999** | **0.999** |
| BA.1 | 0.912 | 0.912 | 0.912 | 0.912 | 0.912 | 0.912 | 0.912 | 0.880 | 0.947 | **0.956** | 0.646 | 0.937 | 0.967 | 0.981 | 0.634 | **0.919** | **0.941** | 0.850 | 0.914 | **0.999** | **0.999** |
| BA.2 | 0.959 | 0.959 | 0.959 | 0.959 | 0.959 | 0.959 | 0.959 | 0.611 | 0.200 | 0.461 | **0.730** | 0.223 | 0.665 | 0.969 | 0.324 | 0.335 | 0.634 | -0.043 | 0.442 | 0.510 | 0.998 |
| Avg. | 0.958 | 0.958 | 0.958 | 0.958 | 0.958 | 0.958 | 0.958 | 0.852 | 0.739 | 0.697 | 0.363 | 0.786 | 0.902 | 0.982 | 0.689 | 0.651 | 0.781 | 0.670 | 0.838 | 0.876 | 0.998 |

**Table 4.**
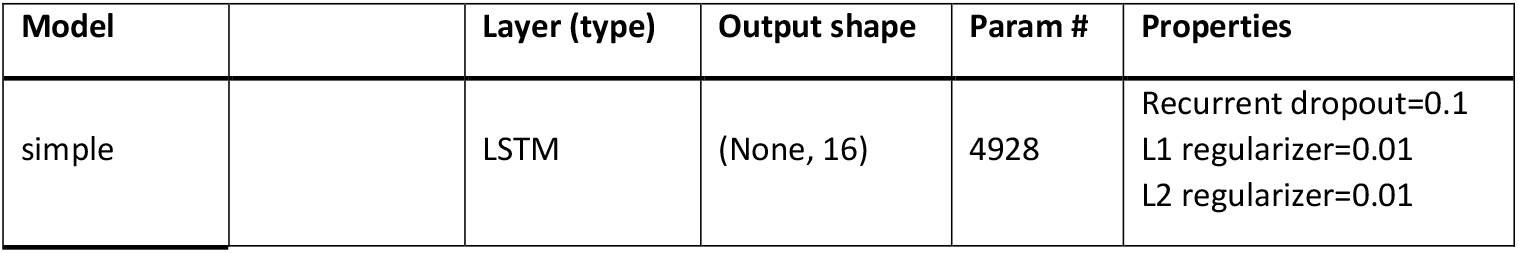
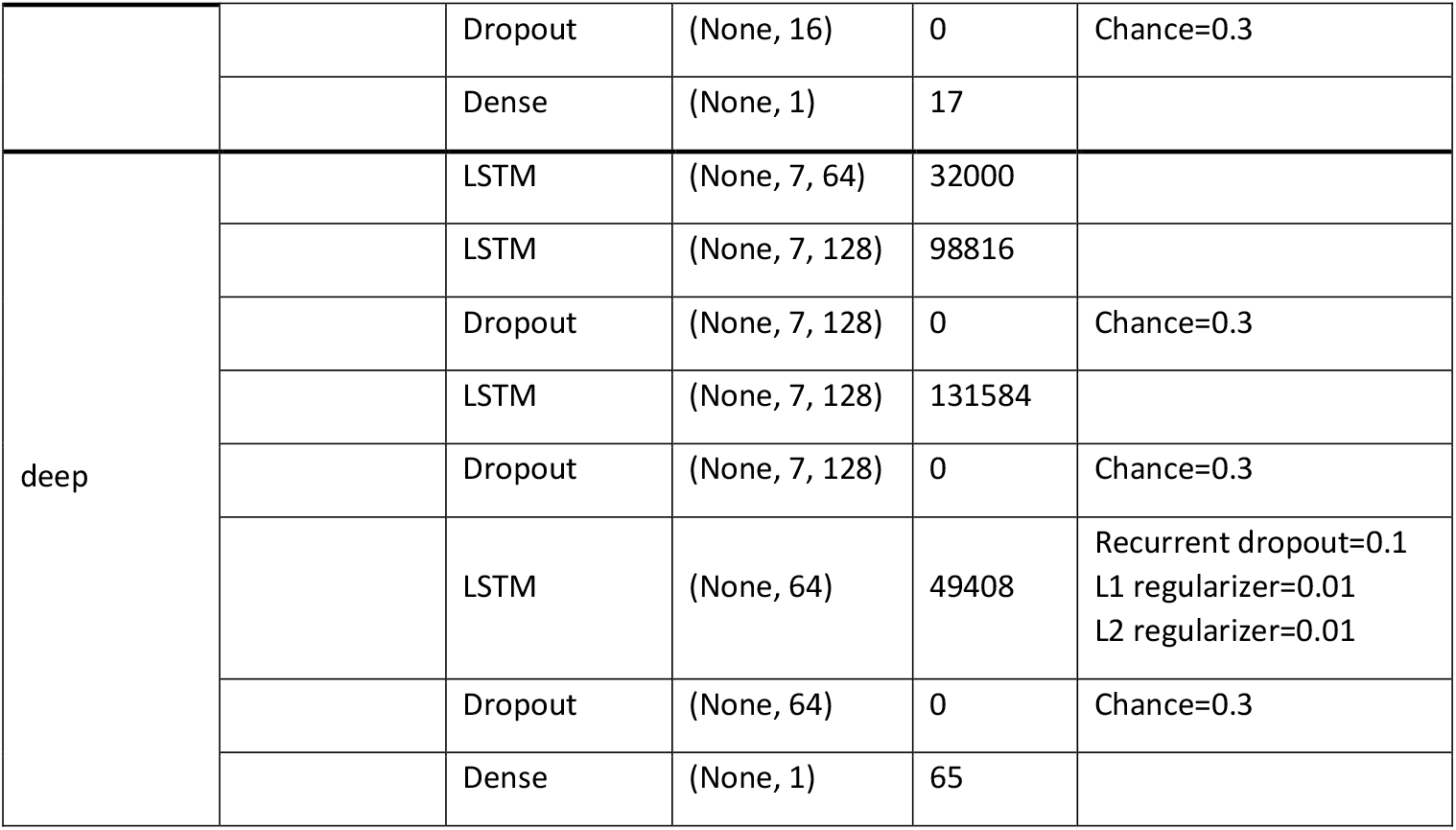
Machine Learning specific details about layer sizes and architecture

## 3 Results

According to the methods described earlier, data was preprocessed, analysed, and evaluated against the reported cases. Error and correlation metrics were used to compare different models and architectures against each other.

### 3.1 Linear Regression

A local linear fit for each of the virus variants (rows) and each of the accumulation periods (columns) with data that has been time shifted already according to maximum Pearson coefficient (Equations (2)-(4)) together with the best linear fit is displayed in Figure 1, where each subtitle also indicates a lag period. A positive lag means that wastewater data leads and case data lags, a negative lag vice versa. Since virus variant contribution varies in time, the input samples for the time lagging, the generation of the Linear Regression model, as well as the calculation of Pearson *r* have been weighted according to the respective variant’s contribution of each sample. This results in a strong influence of samples where a variant is dominant and a weak influence of samples with a relative variant contribution of <0.5. Generally, longer accumulation periods result in a better fit of the Linear Regression and therefore higher values for the Pearson Correlation Coefficient *r* and lower errors. Samples with lower variant contribution (smaller scatter points) do not seem to have a higher likelihood of being outliers in terms of the Linear Model.

**Figure 1.**
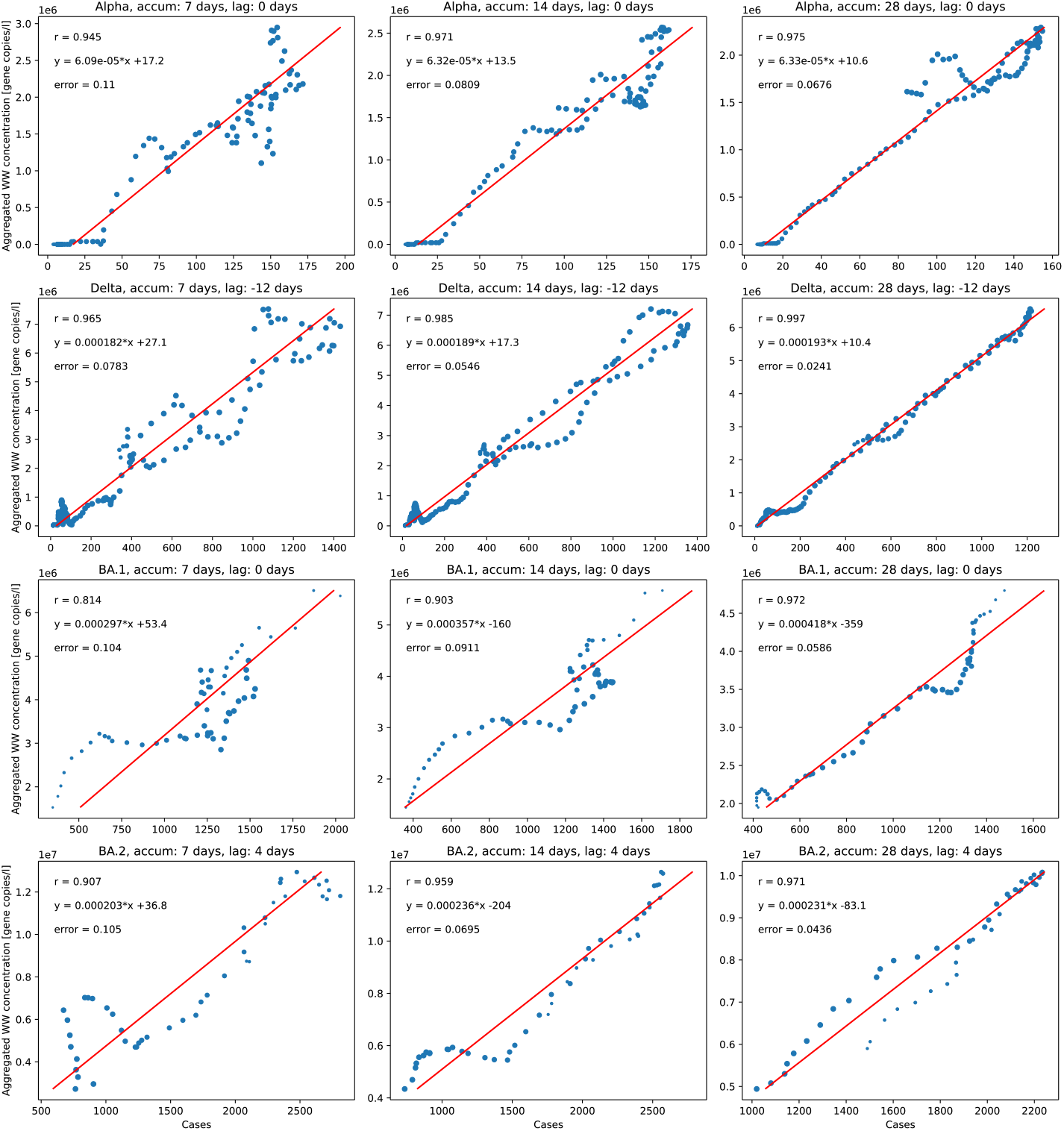
Linear regression for each virus variant and accumulation period (point size relative to strain contribution, r: Pearson correlation coefficient, y: regression equation, error: root mean squared error (RMSE))

Figure 2 shows a combination of all derived linear models. In general, a positive proportional relation between wastewater and case data prevailed. Yet, different curves for each virus variant highlight the differences between the individual models. BA.2 and Delta are very close to the global model, Alpha and BA.1 have a significantly different slope, while all models are roughly pointing towards the origin of coordinates.

**Figure 2.**
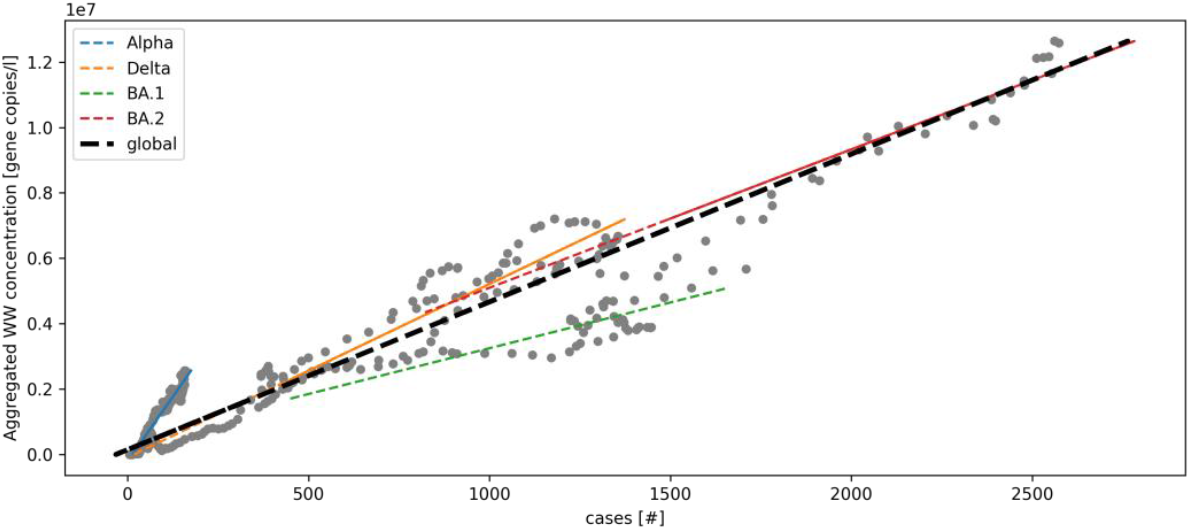
Linear Regression for each virus variant (Blue: Alpha, Orange: Delta, Green: BA.1, Red: BA.2) and combined global linear model (black line). Scatter points: raw data. Accumulated over 14 days.

By applying the regression models to the time series of wastewater data, virus variant specific as well as overall estimates of cases for each aggregation period can be derived (see Figure 3). Each of the models fits the data best in the periods where the respective variant is dominant and is less accurate for other periods. While estimates from the Delta, BA.2 and global model lie very close together, the Alpha and BA.1 models mark the upper and lower boundaries respectively.

**Figure 3.**
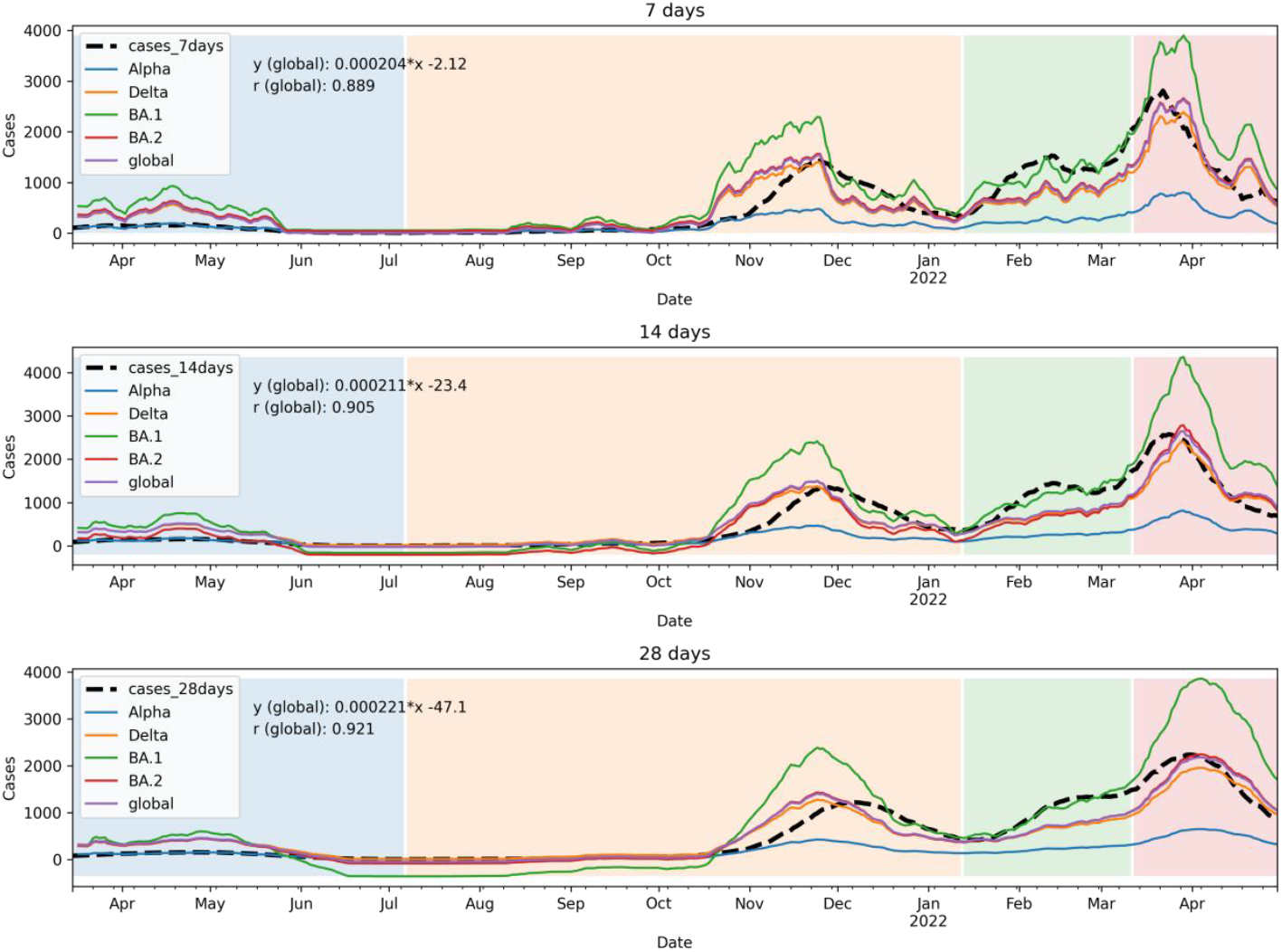
Estimation of reported cases from wastewater data from individual models. Color shades represent local dominance of virus variants. Blue: Alpha, Orange: Delta, Green: BA.1, Red: BA.2

### 3.2 Machine Learning

Machine Learning models have been trained to show an alternative route towards the estimation of actual cases. The input data for this model comprises the wastewater data (accumulated for 14 days only, which poses a good compromise between smoothness and timeliness of the results), virus variant distribution, community vaccination status, precipitation, and finally, as a target vector, the reported cases (14 days accumulated). All input data has been scaled to the target range of 0-1.

After training each model for a maximum of 500 epochs (i.e. iterations of the entire data) until convergence with a batch size of 32 samples (per update of the internal weights), the wastewater data, reported cases, estimate (model output) together with the resulting error are plotted in Figure 4. The red and green shades indicate the model’s variation of estimate and error within ten iterations of the entire training process, including random split of training and validation data. The dashed vertical line indicates the split between training and testing data (between BA.1 & BA.2 wave). Data before the line (2022-03-11) was used for training, while data beyond the line was only used for evaluating the model after training. Here, the BA.2 wave was used as a realistic testing scenario for new variants, which the model will have to predict. While there is good fit of the models in the first three waves, errors increase significantly after that for the last BA.2 wave (dashed line) where they have never seen the training data. The simple model exhibits higher errors in the training section, but shows some correlation in the testing section, where the deep models fails to fit the target curve. Both ML-based models are exhibiting a relatively low variation in the training interval (until BA.2).

**Figure 4.**
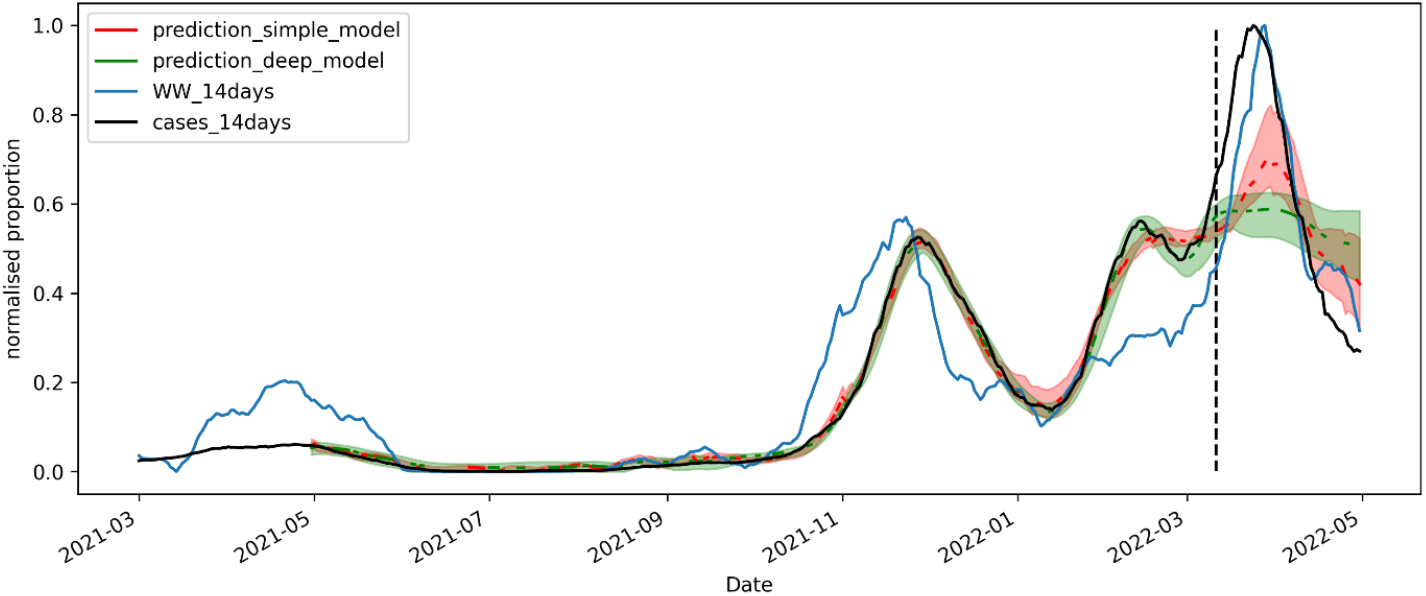
Estimations with the “simple” and “deep” model based on training data up to the BA.1 wave (April 2022). Red and green shades are variations between different training batches.

**Figure 5.**
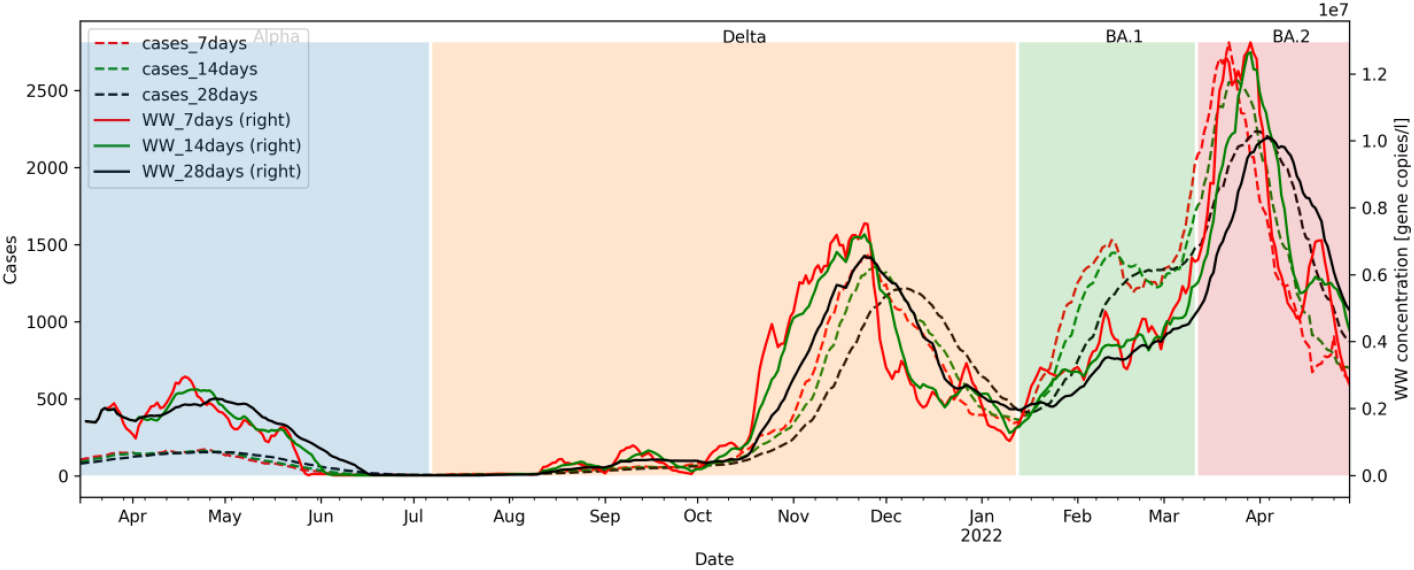
Raw wastewater data and reported cases with different accumulation periods (cases_xdays: reported cases (dashed), WW_xdays: Aggregated wastewater (WW) concentration (each accumulated over x∈[7,14,28] days)). Color shades represent local dominance of virus variants. Blue: Alpha, Orange: Delta, Green: BA.1, Red: BA.2

### 3.3 Comparison of estimation outcome for Linear Regression and Machine Learning

For the comparison of the two methods, the 14-days aggregated input data was segmented according to periods of dominant variants, i.e. Alpha, Delta, BA.1, and BA.2 as shown in the methods section. Models were fitted to the respective individual periods or consecutive multiple periods. Subsequently, the fitted models were applied to the periods of the other variants. For all model-period combinations, after correcting for the time lag of each wave, goodness-of-fit was evaluated separately. Root Mean Squared Errors (RMSE) and Pearson Correlation Coefficients served as indicators for model performance. Results for all models and combinations of training and testing data are displayed in Table 2 and Table 3 respectively. The columns either describe a single wave, for which the model was trained (e.g. “Alpha”) or a series of consecutive waves (e.g. “Alpha->Delta”), i.e. all data included in the period which were used for training.

It can be observed, that the deep ML model, while yielding lower errors on the training data, does not consistently yield lower errors on the test data (Table 2). Variance is low for training and high for testing periods for both ML models (Figure 4). Conversely, the correlation coefficients are high for training and low for testing periods (Table 3).

The Linear Model’s correlation (Table 3) with the training data is lower than that of the ML models, because there is no curve fitting process involved and wastewater data is simply shifted linearly in y-direction. Hence, the correlation coefficient is the same for each wave, independently of the model used, since the time lag is corrected in the evaluation stage of each wave.

Of note, due to the lack of sufficient training data on the Alpha wave (due to 60 days window length), results might not be directly comparable to the linear model for this wave.

## 4 Discussion

While being aware of a multitude of uncertainties in the input data, which are often impossible to quantify (see Limitations), the above work shows that a good correlation can be found when building separate models for each virus variant. In contrast to other findings, those correlations can all be described as linear and none of the axes was transformed. Linearity was inherently assumed at the beginning of the analysis when calculating the time lag. For this calculation, the Pearson coefficient has been used, which is a linear measure. More research could be conducted using non-linear correlation measures, such as spearman or dynamic time warping (Berndt & Clifford, 1994; Spearman, 1961; Tsinaslanidis et al., 2014). Furthermore, potential heteroscedasticity (scale-dependent variance) of the linear correlation models should be investigated.

When analysing Figure 1, some scatter plots showed hysteresis around the linear model which may result from a different time lag depending on whether slopes are rising or falling (e.g.: “BA.2, 28 days”). A potential explanation could be that cases are only reported once when an infection is detected whereas virus material can be found in wastewater for a longer period. This seems to be dependent on the respective virus variant as indicated earlier in the literature, where shedding patterns were found to differ between virus variants (Bertels et al., 2022; Sapoval et al., 2021; Wurtzer et al., 2022). It may be worthwhile to train a separate model (and lag) for each slope of the data to enhance coherence. Different, more fine-tuned aggregation times could be investigated as indicated by some of the literature (Galani et al., 2022; Zhu et al., 2022). Furthermore, different testing efficiencies for establishing case numbers might impact lag time and shape parameters of the functional relation between wastewater and case data (Xiao et al., 2022). A correction of testing efficiency to estimate total infected population, e.g. provided by (Chiu & Ndeffo-Mbah, 2021) might improve the consistency in parameter estimation.

As shown in Figure 3, the different models can be used to estimate today’s rate of new infections for each virus variant. However, as models are derived for individual virus variants, deriving new models for new variants can be very challenging, as case reports may not continue to be as reliable in the future. Potential solutions may be: (1) Using the global model, which is derived from all data, disregarding the influence of different virus variants. This would significantly add to the overall uncertainty but may reduce specific uncertainty for unknown novel virus variants until more information is available. (2) Small scale studies of specific shedding patterns of future variants and their correlation to actual reported cases. This could happen in closed communities, e.g. in hospitals, cruise ships or small towns. Further research would be necessary on how that is applicable to larger WWTPs and areas, such as the case shown. (3) Tune a free modelling parameter for new virus variants, which are easily adjusted as more information about variant specific properties become available. A future model might comprise of e.g. 80% Delta and 20% BA.2 characteristics.

It could be shown that, with relatively little effort, an accurate machine-learning model can be trained for periods where data is available. However, in the testing periods (beyond training period), those models often fail to even correlate to the target variable. Without more insight into the inner workings of the black box or lengthy hyperparameter tuning, it is difficult to build a reasonable predictive model for new data samples, especially new virus variants, as model parameters are difficult to adjust manually. An interesting future research question will be how to update future models with a less than optimal target vector. Still, gaining practitioner’s trust in such models remains a challenge and linear models are often much easier to implement, e.g. in database systems and visualisations. Since the ML models learn from the features in the training data and because each virus variant development is stored in its own feature, new variants are only represented in their own feature, which was not present (or holding meaningful data) during the training process. So whatever characteristics the model has learnt from those features is significantly crippled when they vanish for new testing data. Given that little hyperparameter-tuning has been done there is a large potential for further optimisation of all presented model types and room for research on different model architectures. Furthermore, overfitting might have occurred during the training process and should be investigated.

Given the results of this work, we conclude that ML may not be the best tool for this task because of the mentioned drawbacks and difficult tuning process. Investigating a multivariate Linear Regression is probably a more successful path and is a promising topic for further research.

When case reporting fails to reflect the actual rate of infections, wastewater surveillance can be a surrogate measure, even when observing a slightly higher uncertainty. Wastewater data is easier much less expensive to procure than conducting individual swabs on people, especially on a large scale. When basing pandemic measures on WBE, politics and businesses alike could be using a very robust tool (given further research on uncertainties) that is independent of individual testing motivation, test centre availability or kind of testing procedure (rapid antigen tests vs. PCR). Hospitals could plan their occupation ahead, communities adjust their guidelines and measures and individuals assess their personal risk before meeting anyone. Especially hospitalisation is yet underrepresented in research and should soon be investigated (also see Lünsmann et al., 2022; Zhu et al., 2022).

Furthermore, further research is needed on virus shedding patterns in faeces, viral load concentration and their development over time (Zhu et al., 2022).

## 5 Limitations

In general, due to a lack of standardisation and different population density and different local properties, the results between any two WWTP are not directly comparable. The RKI data source for reported cases only contains locality information on district level (third level territorial units for statistics). Cases are aggregated with a provided district-level code. However, according to the Dresden WWTP, wastewater is collected from twelve municipalities surrounding Dresden area as indicated in the RKI data and amounts for roughly 650k citizens or 613km² (compare Dresden: 556k, 328km²) (Stadtentwässerung Dresden, 2018; Statistisches Bundesamt, 2021). As data for such small entities is not available in the RKI data, it can only be noted as a potential influence factor to explain uncertainties in the model.

### 5.1 Influencing factors on measured virus load in wastewater

As Kitajima et al. (2020) point out, many factors can influence wastewater surveillance data, such as “differences in excretion rates of viruses during the course of infection, temporal delays and the inconsistent capture of spatial variability due to travel and use of multiple wastewater systems in time, and dilution due to precipitation” that may limit virus detection rates. A lack of standardisation of the methods used in each WWTP also makes the results less comparable. A recent comparative study of the recovery rates using three commonly used viral concentration methods found little differences between the methods per se, but highlighted sample turbidity, storage temperature and surfactant load as potential major influence factors in UK wastewater samples (Kevill et al., 2022). Bertels et al. (2022) confirm in their literature review that at least 17 factors can influence measured viral loads in wastewater and try to categorise them, such as shedding-related factors, population size, in-sewer factors, as well as sampling strategy.

Also, the contributions of different virus variants are found to influence the amount of virus material shed into the wastewater through faeces (Weidhaas et al., 2021), specifically, the Omicron (BA.1) variant appears to expose lower levels of virus shedding (low amounts of virus material contained in bodily fluids) (Yuan et al., 2022). It remains an open question how the new BA.2 fits into this. While there are specific PCR, as well as genome sequencing tests available and widely adopted, to detect specific variants of the virus (Wurtzer et al., 2022), it is less relevant for the envisaged correlation of the reported community transmission levels.

As for rain on the sewage systems, Rubio-Acero et al. (2021) found in their data, that precipitation of less than 5mm (in 24h) had no adverse effect on the viral detection performance. They disregarded all days in their data above this threshold and found their data unaffected by rainfall (see also Trottier et al., 2020).

### 5.2 Official case reporting

Governmental strategies for testing and reporting official case numbers vary over time depending on epidemical situation and current political agenda. For example, in the beginning of the pandemic in 2020, when vaccines were not available yet, measures were very strict on a personal and public level. As soon as rapid antigen tests were available, regular testing was demanded by many businesses (e.g. restaurants) as an entry requirement. Upon a positive rapid testing result, citizens were required to get a PCR test that feeds into the official reports. This strategy led to a high percentage of actual discovered infections (and a low number of undetected infections). An additional direct influence factor on reported cases refers to the varying access to testing (Olesen et al., 2021).

The more vaccinations had been administered and hospitalisation rates went down, the less often people had to get themselves tested. Furthermore, some virus variants (e.g. Omicron) cause milder symptoms, which often do not require medical treatment, which leads to more people just staying home instead of going to a test centre (Maisa et al., 2022). On public holidays and in times of high community transmission rates, it can be very challenging to find free capacities in either a test centre or a laboratory or even get the results back on time.

All of the earlier mentioned uncertainties in the input data might explain any outliers and diversions, especially in the linear model. With the current knowledge, it is near impossible to quantify any of those effects so that the only option at this point is to accept them as the given approximation.

### 5.3 Computational shortcomings

Due to the focus of this work and partly due to the lack of computational resources, intensive hyperparameter tuning has not been done on the deep learning models. It is expected that the efficacy of the models could be enhanced with proper fine-tuning but was simply out of scope in this work. Different model sizes have not been considered systematically.

## 6 Conclusion

The use of wastewater-based epidemiology has been evaluated in light of the current SARS-CoV-2 pandemic. Input data has been procured and analysed. Virus variant based Linear Regression models and ML models have been built, and their performance compared. Lastly, their use cases, shortcomings and potential fields of further research have been discussed.

We hereby conclude that quantified virus particles in the wastewater can be a reasonable approximation for the reported cases. However, the accuracy and detail of the modelled correlation is highly virus variant dependent, which is why a continuous adaptation of the currently most dominant variant in terms of availability and verifiability is essential for our proposed approach. Since it is rather difficult to obtain precise information about the dilution of virus particles of new virus variants, one may use information from other local areas with newly arising variants and parametrize the new model accordingly. The respective variant information could be tested via qPCR or sequencing approaches. In turn, we suggest the incorporation of virus variant information into other related forecasting models, which can be an essential asset because it addresses different existing and novel variants at the same time.

The proposed linear model can easily be incorporated into any database or visualisation platform, as well as parameterised and extended. It can be of great use for any authority, clinical, and political decision makers, as well as the public as an indication of the current level of local infections.

## Data Availability

All used scripts and raw data are available online at

https://github.com/MM-Lehmann/Wastewater-modelling-Dresden

## 8 Acknowledgements

This work was in part supported by a grant of the State Ministry of Science and Cultural Affairs of Saxony (SMWK). The authors thank the employees of the wastewater treatment plant Dresden for their collaboration and Miss Dipl.Biol. Sara Schubert for the preparation of the ethics application.

## 9 Author contributions

M. Lehmann was responsible for the study design; M. Geissler, R. Dumke, and A. Dalpke conducted the experiments; M. Lehmann, W. Hahn, R. Gebler, B. Helm and M. Wolfien analysed and interpreted the data; M. Lehmann and W. Hahn were involved in image generation; M. Lehmann drafted the first version of the manuscript; M. Lehmann, M. Geissler, W. Hahn, R. Dumke, A. Dalpke, and M. Wolfien carefully revised the final version of the manuscript. All authors read and approved the final version of the manuscript.

## 10 Conflict of Interest

The authors declare that no conflict of interest exists.

## 11 Ethical considerations

Ethics application (BO-EK-383072021) has been confirmed by the Ethic Committee of Technische Universität Dresden, which is registered as institutional review board (IRB00001473) at the Office of Human Research Protection.

## 12 Data Availability

All used scripts and raw data are available online at https://github.com/MM-Lehmann/Wastewater-modelling-Dresden publicly.

## 13 Supplements - ML model architecture

## Notes

### Competing Interest Statement

The authors have declared no competing interest.

### Author Declarations

Ethics application (BO-EK-383072021) has been confirmed by the Ethic Committee of the Dresden Technical Univsersity, which is registered as institutional review board (IRB00001473) at the Office of Human Research Protection.

